# Chest X-ray image analysis and classification for COVID-19 pneumonia detection using Deep CNN

**DOI:** 10.1101/2020.08.20.20178913

**Authors:** Terry Gao

## Abstract

To speed up the discovery of COVID-19 disease mechanisms by X-ray images, this research developed a new diagnosis platform using a deep convolutional neural network (CNN) that is able to assist radiologists with diagnosis by distinguishing COVID-19 pneumonia from non-COVID-19 pneumonia in patients based on chest X-ray classification and analysis. Such a tool can save time in interpreting chest X-rays and increase the accuracy and thereby enhance our medical capacity for the detection and diagnosis of COVID-19. The research idea is that a set of X-ray medical lung images (which include normal, infected by bacteria, infected by virus including COVID-19) are used to train a deep CNN that can distinguish between the noise and the useful information and then uses this training to interpret new images by recognizing patterns that indicate certain diseases such as coronavirus infection in the individual images. The supervised learning method is used as the process of learning from the training dataset and can be thought of as a doctor supervising the learning process. It becomes more accurate as the number of analyzed images grows, and the average accuracy is above 95%. In this way, it imitates the training for a doctor, but the theory is that since it is capable of learning from a far larger set of images than any human, it can have the potential of being more accurate.

## 1 Introduction

Coronaviruses are a large family of viruses that cause illness ranging from the common cold to more severe diseases, such as Middle East Respiratory Syndrome (MERS-CoV) and Severe Acute Respiratory Syndrome (SARS-CoV) [1]. This novel coronavirus (COVID-2019) [2][3][4] is a new strain not previously identified in humans. A common clinical feature of severe COVID-19 infection is pneumonia [5][6][7][8]. Chest X-rays are a useful diagnostic tool for assessing various lung diseases, such as pneumonia, but interpretation of the images can be challenging and time consuming [9] [10].

Part of the challenge is distinguishing between normal tissue and disease processes, a skill that must be learned through experience, particularly for some illnesses such as pneumonia where the difference is less obvious. With a great number of patients having chest X-rays taken as part of the diagnostic examination of suspected pneumonia each year at hospital alone, the evaluation of X-rays consumes a considerable amount of resources.

Machine learning technology is currently being implemented in a variety of different fields, including diagnostics and bioinformatics. A convolutional neural network (CNN) is a deep learning algorithm [11] that can be implemented in medical image processing to support correct and speedy decision making [12][13][14][15]. The general idea is that a set of medical images is used to train a deep learning CNN that is able to distinguish between noise and useful diagnostic information [16][17][18][19]. The CNN then uses this training to interpret new images by recognizing patterns that indicate certain diseases in the individual images. In this way, it imitates the training of a doctor, but the theory is that since it is capable of learning from a far larger set of images than any human, the CNN approach has more accurate results.

A pilot study using publicly available chest X-rays of no pneumonia patients and patients with coronavirus should promise in that it is possible to train a CNN to distinguish between these two groups with approximately 90% high accuracy [20]. Although this pilot cannot be translated directly to an actual clinical situation, as the analysis is based on digital image processing, it is possible to train a CNN to assist medical staff in distinguishing pneumonia from no pneumonia patients based on X-rays. In addition, there is the potential to distinguish viral from bacterial pneumonia, which is particularly relevant to COVID-19 infection because pneumonia is directly associated with the virus rather than a bacterial complication. The research is that a set of X-ray medical lung images used to train a deep CNN can distinguish between noise and useful information and then uses this training to interpret new images by recognizing patterns that indicate certain diseases such as coronavirus infection in individual images. The supervised learning method is used as the process of learning from the training dataset and can be thought of as a doctor supervising the learning process. It can be more accurate as the number of analyzed images grows. Such a tool could increase the speed and accuracy of interpreting and thereby improve the overall treatment of patients, which is useful for COVID-19 disease detection.

The product developed by this research can reduce the workload of doctors and detect disease at the early stage, so it has precisely customers and scheduled finished as soon as possible. In the future, I can also use the similar system developed in this research to show signature patterns in other medical image data, such as CT, MRI, MEG, etc. This research can range from lung disease detection to heart disease or cancer detection, which can help to change how I treat early diagnosis.

## 2 Design and Methods

If X-rays are used as a means for COVID-19 diagnosis, we need to take into account that diagnostic decisions are made on individuals, not on groups. Due to the limited health care resources in New Zealand, a fast decision system can help doctors automatically process individual data and make accurate predictions of the possibility for one to develop a particular disease; this requires a fast and personal specific health system, but the current system for pneumonia detection primarily relies on humans, which have reasonable accuracy but with a high cost of time and resources. The average time it takes a trained radiologist to read a chest X-ray image is approximately 5-6 minutes. It is difficult to speed that up because chest X-ray reading is a very systematic process. Additionally, training enough radiologists for growing demands within a short period of time is impractical, if not impossible. This research involves some technological breakthroughs, where artificial intelligence, such as deep learning and machine learning methods, comes into play and helps to improve the diagnostic efficiency in New Zealand.

### 2.1 Study Design and Type

This research built a diagnostic system that uses open-to-public coronavirus infector chest X-ray images from [21] for training. The historical data are split into a training and a validation set. The CNN is then trained on the training set and the predictive value of the tool, once trained, and determined by using the validation set. Tests of what parts of the images by which the CNN uses to determine the output are explored to ensure that the output is clinically relevant. After this initial analysis, a massive extraction of texture features was applied and can serve to provide additional information for the diagnosis of COVID-19.

### 2.2 Participants

The training of the CNN needs to have no pneumonia and pneumonia representing X-rays that are alike in all other aspects that may influence how an X-ray looks, so the deep CNN is trained to look at the actual difference based on the presence of pneumonia and no other factors associated with pneumonia. Since a patient who is diagnosed and treated in-house has at least an X-ray to diagnose the condition and an X-ray to confirm that the pneumonia is gone, we have X-rays from the same patient with and without pneumonia. For this reason, the training set is a random selection of patients. The chest X-rays used are the first X-ray taken of the patient during the admission from the moment a pneumonia was suspected, and the last X-ray taken before discharge. Both X-rays taken used the same position of the patient (standing/lying in bed).

### 2.3 Outcomes

For each record in the validation set, the following outcomes Ire collected:

- Diagnosis as determined by the trained CNN
- Gold standard diagnosis, as determined by radiologist and confirmed by discharge ICD10 codes.
  ∘ If an X-ray as determined not to show pneumonia but discharge ICD10 code shows pneumonia, then at least one other X-ray of that admission episode has to have shown pneumonia and the X-ray cannot be flanked by two X-rays showing pneumonia. If either of these two criteria is not met, then the X-ray is determined by showing pneumonia.
  ∘ If an X-ray was determined to show pneumonia, then the discharge ICD10 code had to show pneumonia; otherwise, it was determined not to show pneumonia.

### 2.4 Sample size calculations

The training data size depends on the complexity of the CNN model, such as the number of inputs/outputs, the relationships between parameters, the noise in the data, and the variance and standard deviation of every parameter, so the best approach is to ensure that our data cover all the ranges we want for all parameters. Normally, the number of samples is at least 10 times more than the number of CNN training parameters, so we initially set the training samples to approximately 1400 chest X-ray images, which include 400 normal images, 400 pneumonia infected by bacteria images, 400 pneumonia infected by other virus images, and 200 pneumonia infected by COVID-19 images. Testing samples are approximately 400 chest X-ray images (100 images for each class). Approximately 100 chest X-ray images are used for validation, which also include approximately 50 COVID-19 infection images.

## 3 Implementation Procedure

The primary step of this research is a deep CNN designed and trained to assist radiologists with diagnosis by distinguishing COVID-19 pneumonia from non-COVID-19 pneumonia in patients at hospital with high predictive values using clinically relevant parts of the images. Then, this deep CNN is used to distinguish bacterial from viral pneumonia amongst those patients with pneumonia at hospital with high predictive values using clinically relevant parts of the images. I designed the CNN based on VGG-19 in Fig. 1 and reduced the levels, changing the convolutional kernels to make it more feasible.

**Fig. 1.**
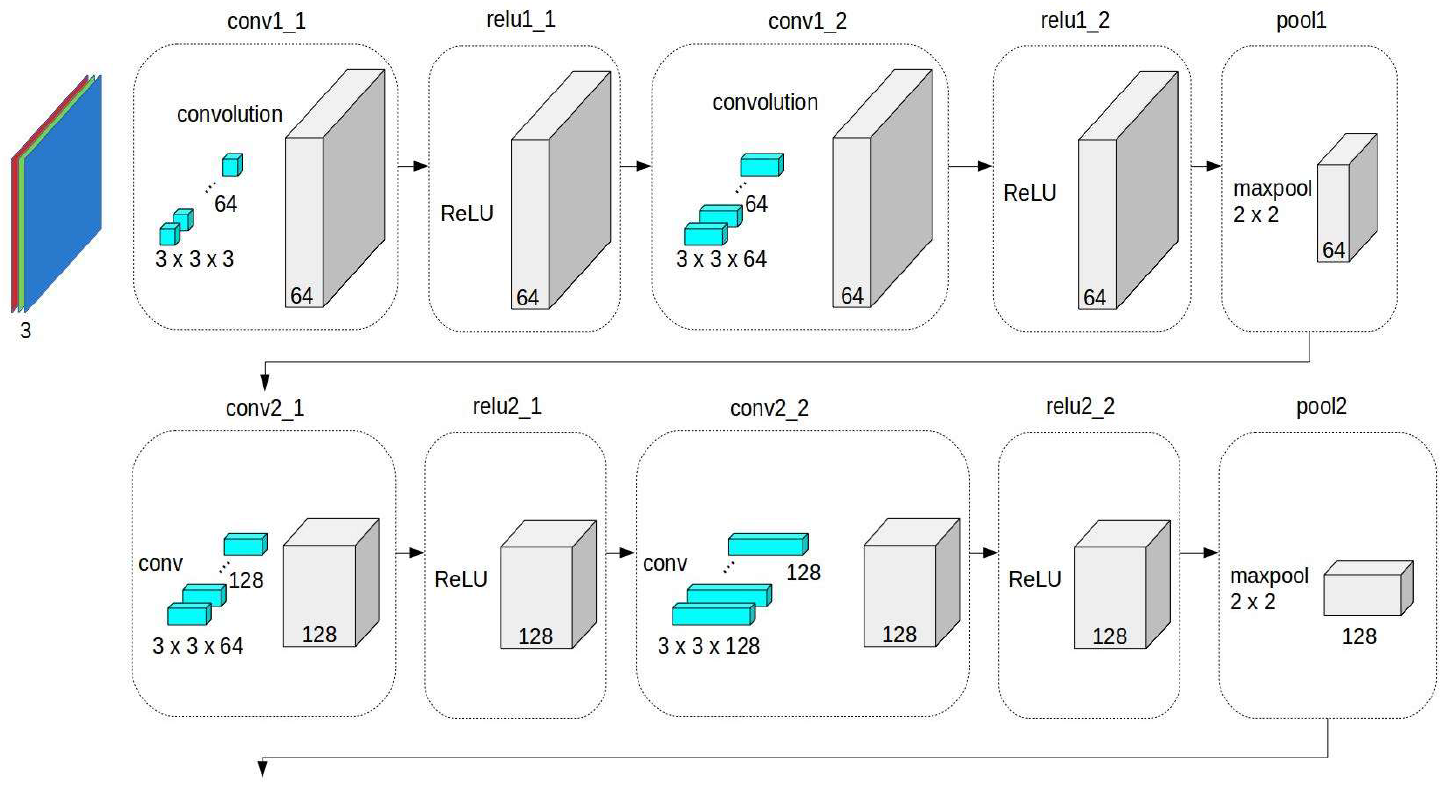
VGG-19.

Let x be set as the input vector, *φ*_*i*_ is the radial basis function, *N* is the number of input training samples, and *y* is the output of the neural network:

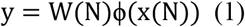

Then, *d* (*n*) is the output response of the *n* iterations of the neural network, and the error is defined as:

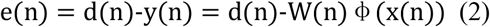

and the objective function as:

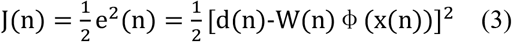

and to the weight is updated according to:

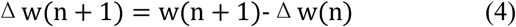

According to the gradient descent with the momentum algorithm, I can obtain:

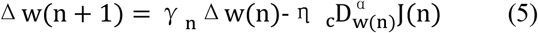

where *η* is the learning rate, 0 < *α* < 1, 0 < *γ* < *η, γ* is the momentum factor, and *γ*_*n*_ is the momentum coefficient designed as follows:

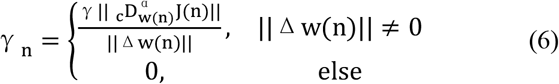

||·|| is the Euclidean normalization. According to the definition of the fractional derivative, I can obtain the final result as:

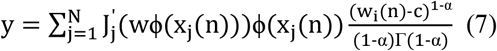

### 3.1 Training the CNN

The convolution layers have a hierarchical structure and are core building blocks of a CNN. Fig. 2 depicts the structure of the CNN in this research by using the visualization tool [23]. Deep CNNs applying individual network levels and rapid combinations of features take place before the forecasting stage. The input of the first convolution layer is the input space, and the output is the feature map. The input and output of the next convolutional layers are feature maps of the input space. The number of convolutional layers is set by the programmer. The set of feature maps is obtained as the output of convolutional layers. The complex features of the input space are represented by using the stacked hierarchical structure of convolutional layers. The obtained features from the convolutional layers are fed to the pooling layer. An activation function such as ReLU is applied to the obtained feature map. In this layer, the relevant features are retained, and the rest are discarded. A dropout layer with a dropout factor of 0.5 has also been used for the regularization of the model. Then, the feature maps of the corresponding depths of the contraction path are fed as input. The obtained features are transformed into a one-dimensional array called the feature vector. The feature vector is a one-dimensional array and is the input for the fully connected layer. The fully connected layer calculates the output of the CNN.

**Fig. 2.**
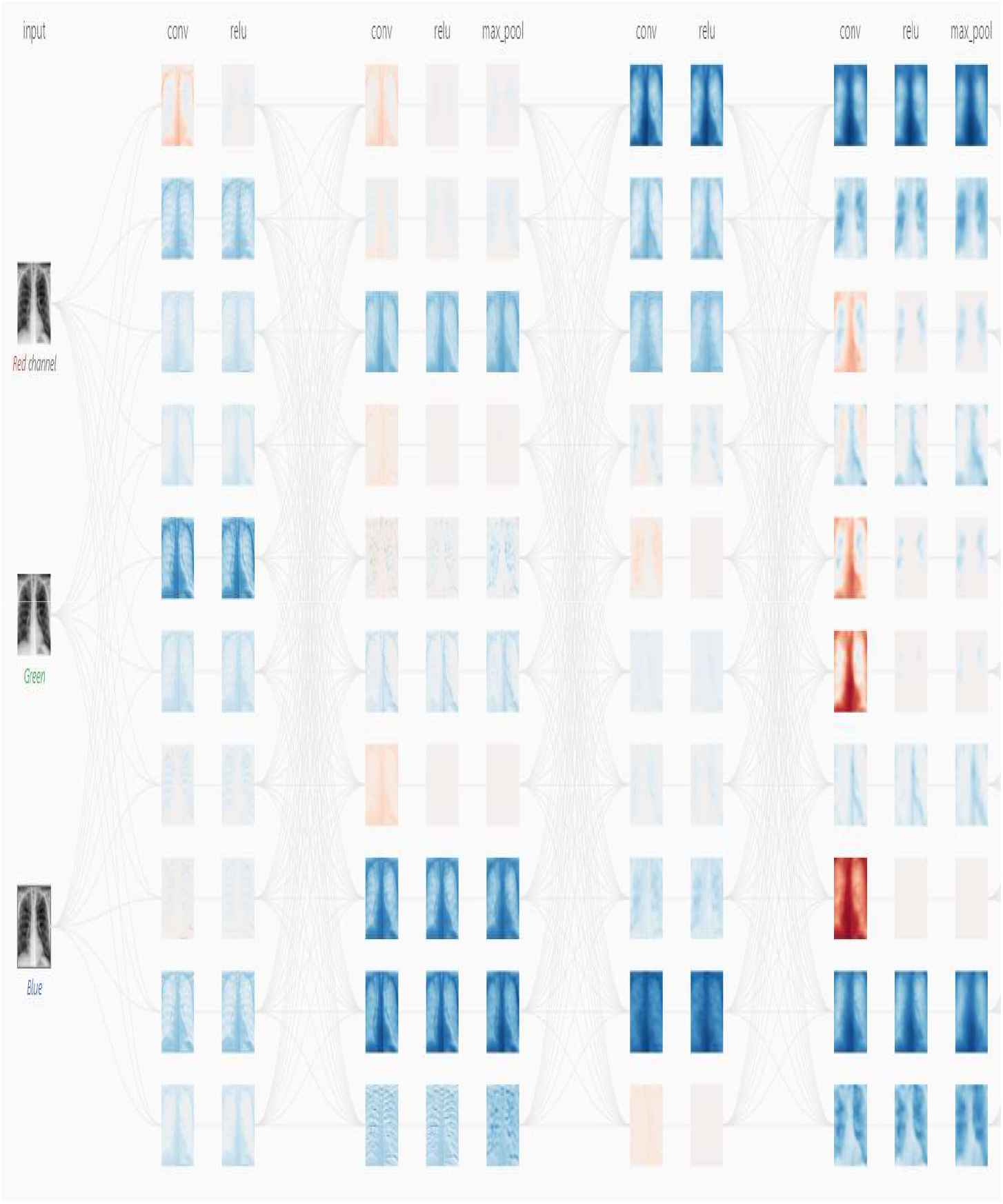
The structure of CNN in this research.

The regression branch predicts the distances from the center of each grid to the four sides of the bounding box. Centeredness is a coefficient in the range of [0,1] for each grid. The farther the grid center is from the object center, the smaller the coefficient is. The centeredness and class are multiplied and then serve as the input of nonmaximal suppression (NMS). The sclera block is similar to a fully convolutional network (FCN), where the input feature map is unsampled 4 times to obtain a score map. After these operations using the ReLU activation function, the nonlinear transformation of signals is performed for each matrix. The obtained results are sent to the pooling layer. In this layer for each cell, the max (or average) pooling operations are performed. In the pooling layer, down sampling operations have been performed to reduce the size of the feature matrix derived from the convolution layer. After training, the class index is used to measure the class activation map, and the layers can be used when visualizing the class activation map. The CNN gradient model is constructed by supplying the inputs of the pretrained model and the output of the final layer in the network. The average of the gradient values is computed by using connection Weights, and the ponderation of the filters is computed with respect to the Weights, so the connection heatmap can be formed and normalized such that all values are set in the range [0, 1], and the resulting values can be scaled to the range [0, 255] to finally show the regions of interest with bright color that can be used for medical purpose analysis. Details are as in Table 1.

**Table 1.** The parameters of each level for CNN.

|  | Layer Type | Output Shape | Param |
| --- | --- | --- | --- |
| Input Layer | Input | 64 x 64 x 3 | 0 |
| Hidden Layer 1 | Conv1 | 64 x 64 x 32 | 896 |
|  | ReLU | 64 x 64 x 32 | 0 |
|  | Pool1 | 32 x 32 x 32 | 0 |
| Hidden Layer 2 | Conv2 | 32 x 32 x 64 | 18496 |
|  | ReLU | 32 x 32 x 64 | 0 |
|  | Pool2 | 16 x 16 x 64 | 0 |
| Hidden Layer 3 | Conv3 | 16 x 16 x 128 | 73856 |
|  | ReLU | 16 x 16 x 128 | 0 |
|  | Pool3 | 8 x 8 x 128 | 0 |
| Classification layer | Flatten | 8192 | 0 |
|  | Dense1 | 16 | 131088 |
|  | ReLU | 16 | 0 |
|  | Dense2 | 64 | 1088 |
|  | ReLU | 64 | 0 |
|  | Dense3 | 128 | 8320 |
|  | ReLU | 128 | 0 |
|  | Dense4 | 2 | 258 |

The first convolutional layer learns 32 convolutional filters, each of which is 3×3. Then, rectified linear units (ReLUs) are applied as an activation function that has output 0 if the input is less than 0 and output otherwise. The following layers use similar processing. The fully connected layer uses SoftMax for the activation function.

### 3.2 Testing the Deep CNN

Once the CNN has been trained using the training set, it is used to diagnose all the X-rays in the test set. For each case, the proportion of each diagnosis can be obtained. The parameters used to indicate the performance of CNN are as shown in Fig. 3 below:

**Fig. 3.**
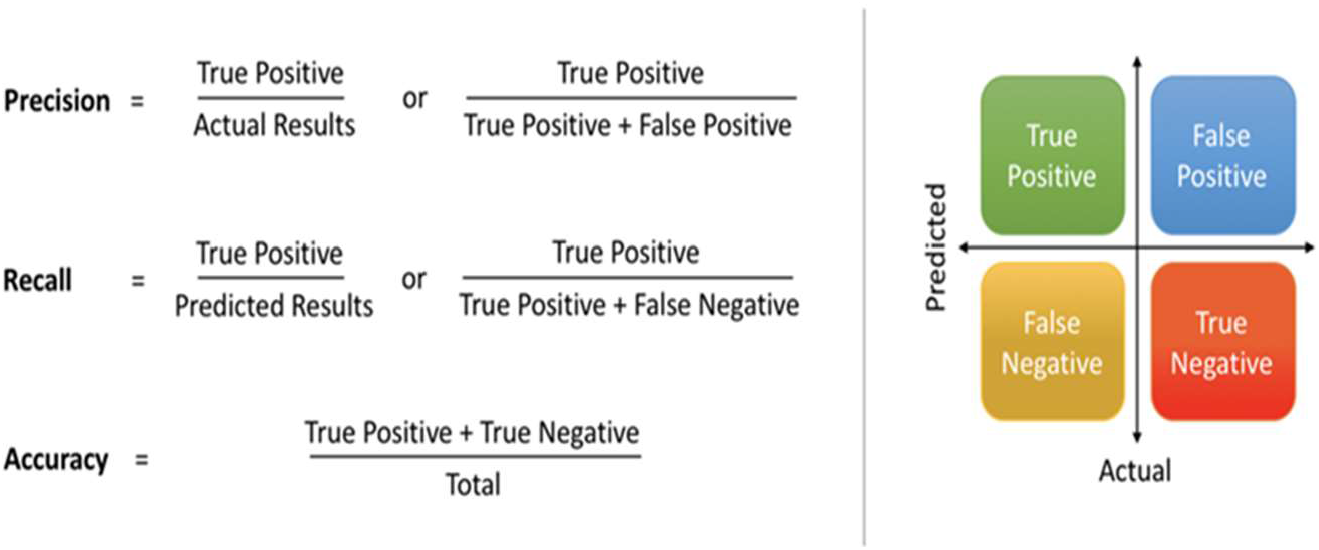
Accuracy parameters.

### 3.3 Experimental Tests

The testing rules used are as follows:

1. Positive and predictive values are determined with family-wide 95% confidence intervals (Bonferroni correction) for the output of the CNN in determining COVID-19 pneumonia.
2. Positive and predictive values are determined with family-wide 95% confidence intervals (Bonferroni correction) for the output of the CNN in determining bacterial against viral pneumonia.

An example of the type of X-ray data that is analyzed using our CNN approach is shown in Figure 1. The test image is from “The New England Journal of Medicine, 2020: January 31. DOI: 10.1056/NEJMoa 2001191”. [22]. The data augmentation methods applied in the proposed CNN are scale, shift, rotate, salt and pepper noise, and flip. By applying these small transformations to images during training, variety in the training dataset has been created and improves the robustness of the proposed model. Generators are implemented for dynamic augmentation of the input image and generation of the corresponding ground truth labels.

The experiment and software are based on TensorFlow 2.1-GPU, Python 3.7 and CUDA 10.1 for accelerated training. The hardware for this system includes two i7-CPUs, 16.0 GB memory, and a 500 GB SSD drive, NVIDIA GeForce GTX 1660 Ti GPU, and it takes approximately 4 hours of training to converge. The user interface of the system developed in this research is shown in Fig. 4 and Fig. 5.

**Fig. 4.**
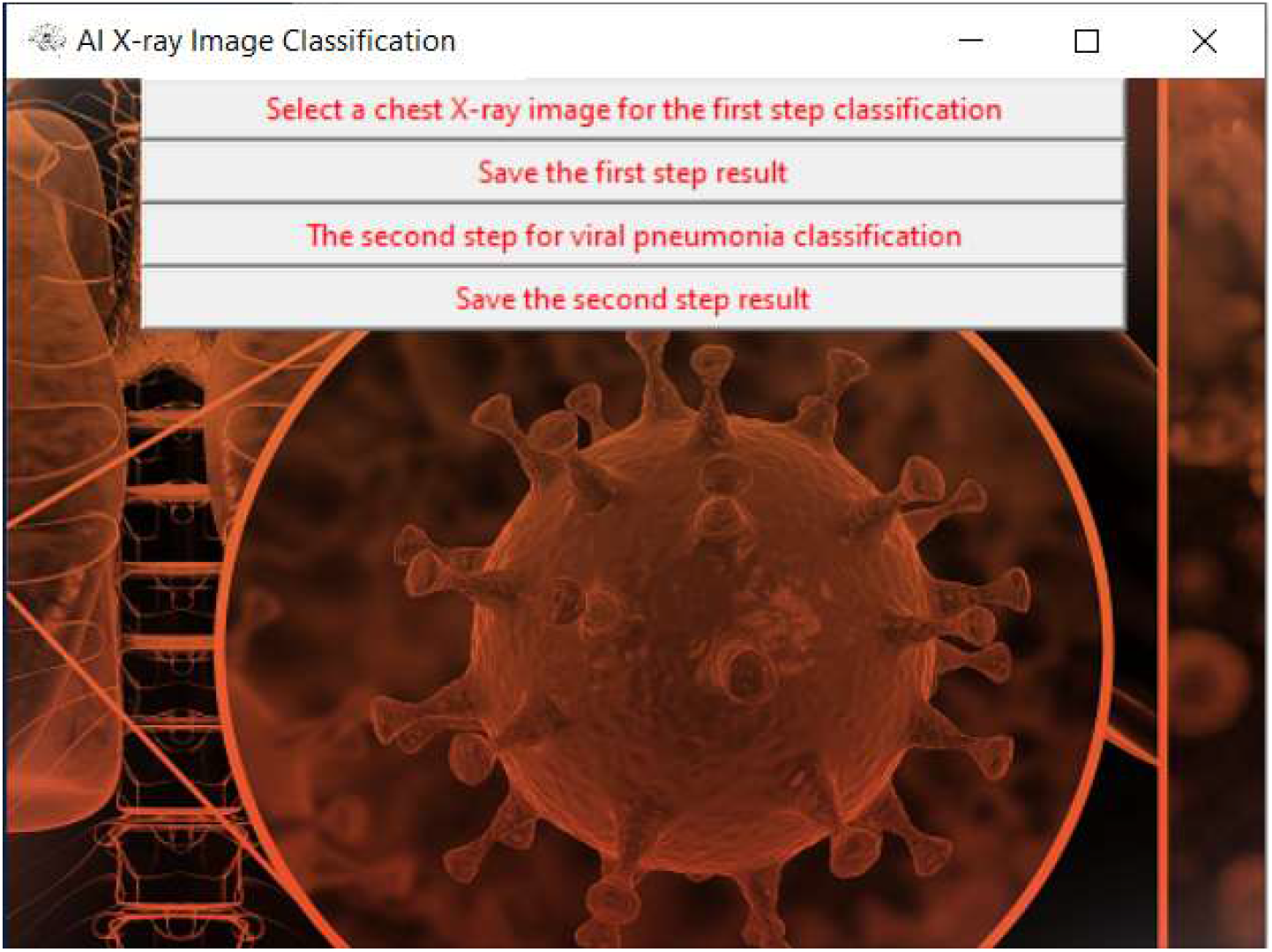
User interface (UI) of the system.

**Fig. 5.**
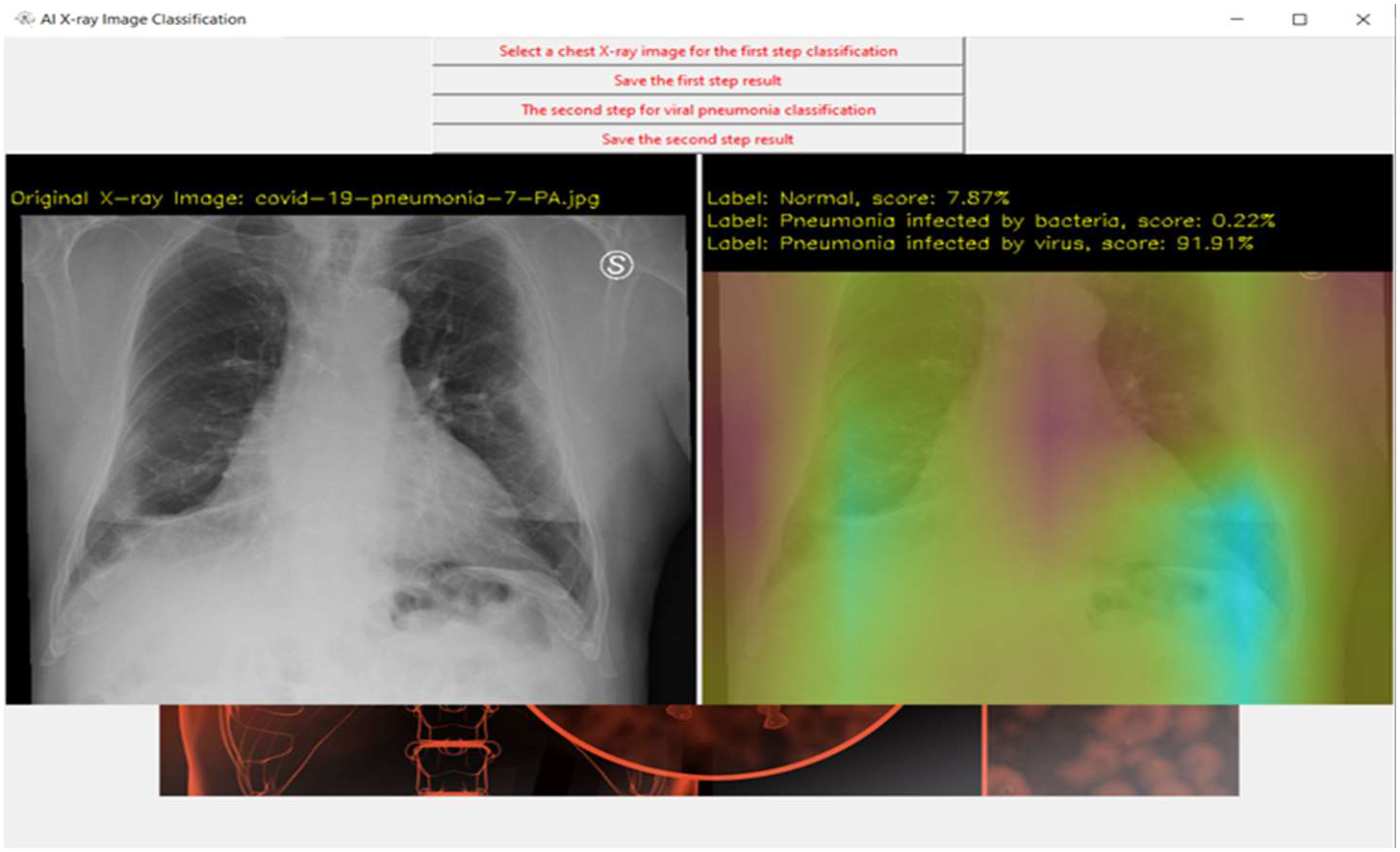
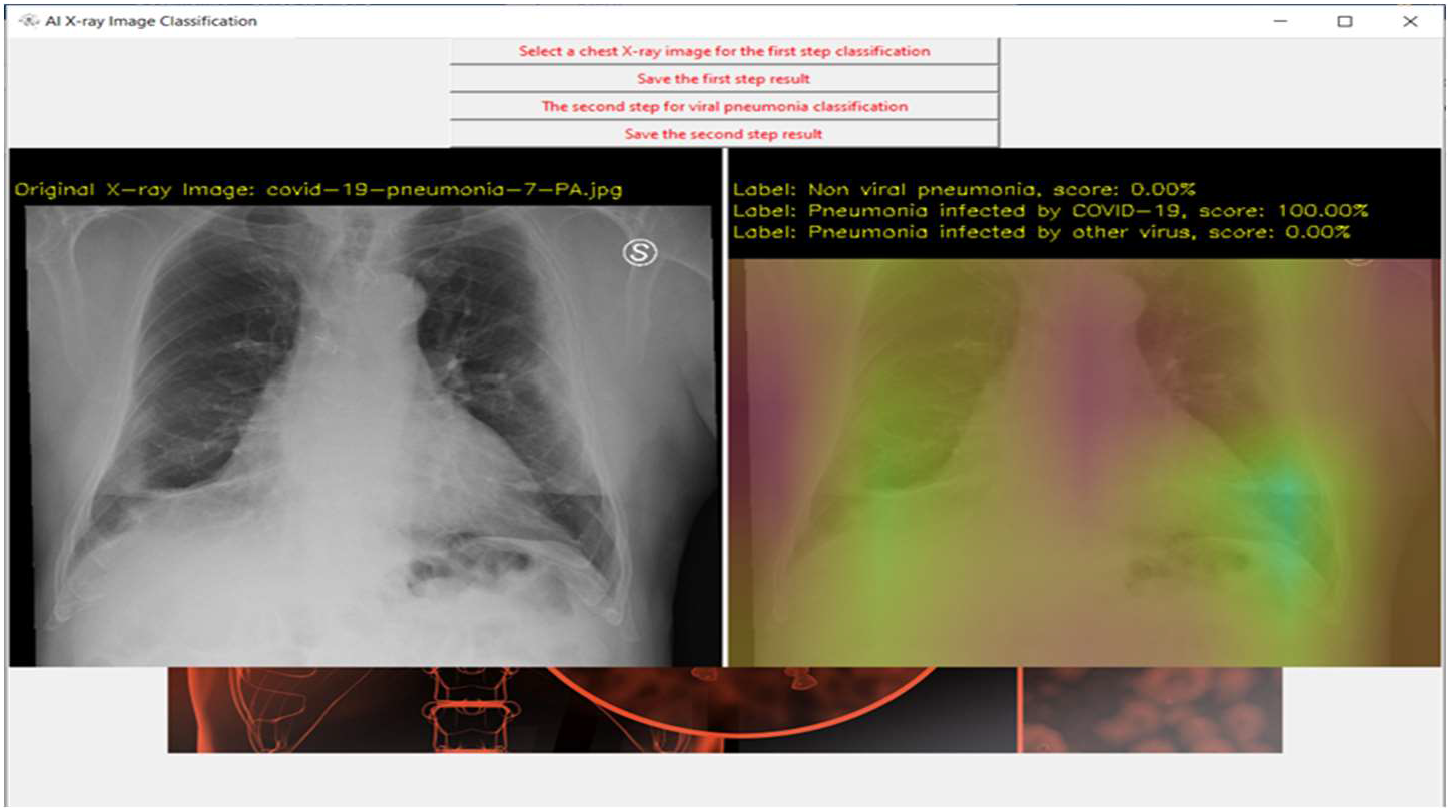
Analysis Results Demo. Fig. 5-a) Results from step 1. The left image shows the original X-ray data, the right image shows the possibility of 3 different cases (normal, infected by bacteria, infected by virus), and the brighter color shows which part has the potential infection problem. Fig. 5-b) Results from step 2. If step 1 shows viral pneumonia with the highest score, continuous processes are analyzed and checked if the patient has COVID-19 infection or not. The right image shows the possibility of 3 different cases (non-viral pneumonia, infected by COVID-19, infected by other viruses), and a brighter color shows which part has the potential infection problem.

The architecture of CNN categorizes benefits in X-ray medical imaging, such as the number of modules in interconnected operations and input modalities, dimension in input patch, quantity of time predictions and contextual information about implicit and explicit. The test results of the proposed CNN are shown in Table. 2 and Fig. 6.

**Tab.2.** Experimental test results.

| Parameters |  |  |  |  |
| --- | --- | --- | --- | --- |
| X-ray Category | Precision | Recall | F1-score | support |
| **Normal** | 0.97 | 0.94 | 0.95 | 572 |
| **Pneumonia infected**<br>**by Bacteria** | 0.99 | 0.93 | 0.96 | 213 |
| **Pneumonia infected**<br>**by COVID-19** | 0.95 | 0.98 | 0.97 | 117 |
| **Pneumonia infected**<br>**by normal Virus** | 0.98 | 0.96 | 0.97 | 410 |
| **Average** | 0.97 | 0.95 | 0.96 | 1252 |

**Fig. 6.**
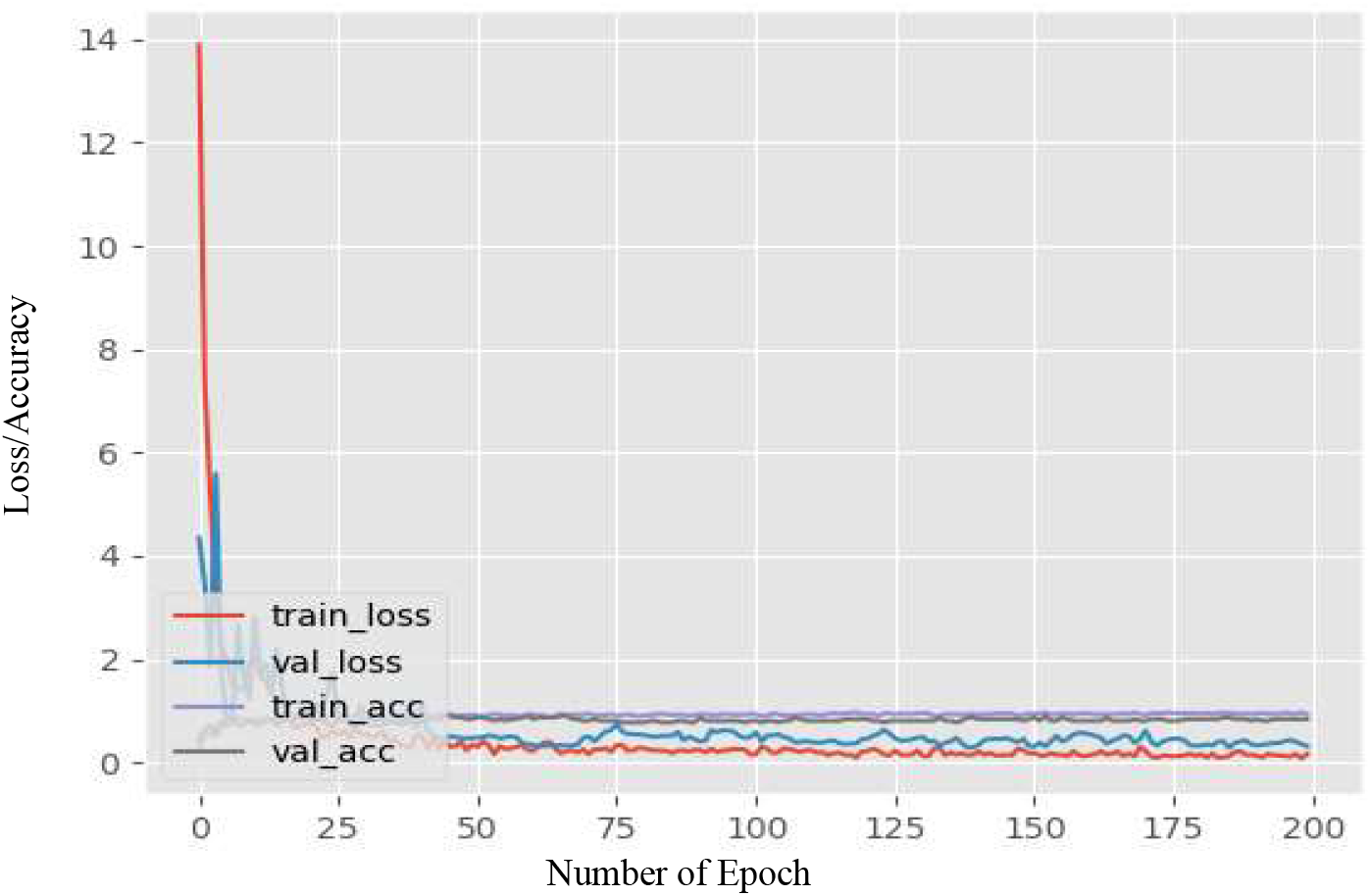
Loss and Accuracy.

## 4 Conclusions

X-ray images play an important role in the diagnosis of COVID-19 infection from other pneumonia as advanced imaging evidence. Artificial intelligence (AI) algorithms and radionic features derived from chest X-rays can be of great help to undertake massive screening programs that could take place in any hospital with access to X-ray equipment and aid in the diagnosis of COVID-19. As all the process can be done automatically, the cost is significantly decreased compared with traditional methods. To speed up the discovery of disease mechanisms, this research developed a deep CNN-based chest X-ray classifier to detect abnormalities and extract textural features of the altered lung parenchyma that can be related to specific signatures of the COVID-19 virus. In this way, it imitates the training for a doctor, but the theory is that since it is capable of learning from a far larger set of images than any human, it has the advantages of being more accurate and reducing the infection diagnosing time.

## Data Availability

https://www.kaggle.com/paultimothymooney/chest-xray-pneumonia

